# Dynamics of the Third wave, modelling COVID-19 pandemic with an outlook towards India

**DOI:** 10.1101/2021.08.17.21262193

**Authors:** Ayanava Basak, Sayanur Rahaman, Abhishek Guha, Tanmay Sanyal

## Abstract

Since 2020, the COVID-19 pandemic has devastated human civilization throughout the earth. The pandemic is returning in different waves because of constant changes in the genetic components of the virus. Had we been able to predict the nature and timing of these waves earlier, numerous lives could, in essence, have been saved. It is evident that the situation has spiraled out of control in several countries for want of proper preventive measures. In this article, we described a comprehensive mathematical approach to understand the nature of the pandemic waves. Also, we determined the probable timing of the third wave that will help the concerned government(s) to take the necessary steps to better prepare for the unforeseen situation.

## 1. Introduction

The entire world is passing through a difficult situation because of coronavirus disease-19 (COVID-19) outbreak since January 2020. Severe acute respiratory syndrome coronavirus 2 (SARS-CoV-2) was identified as the cause of this disease. All human corona viruses (HCoVs) are zoonotic and contain RNA as genetic material. Before the outbreak of Severe Acute Respiratory Syndrome (SARS) and Middle East Respiratory Syndrome (MERS), HCoVs were known to cause mild illness. The SARS (in 2003) and MERS (in 2012) for the first time, showed how devastating and life-threatening an HCoV infection could be. The emergence of SARS-CoV2 a in the Wuhan province of the People’s Republic of China at the end of December 2019 has brought the CoVs back into the limelight again^[1-4]^. As of May 2021, COVID-19 has affected 222 countries, changing the daily lives of billions of people^[13]^. It is a huge challenge for the doctors and scientists to save the mankind from this pandemic. Although the first wave of the pandemic progressed slowly in many countries, the second wave has been more severe and devastating. Unfortunately, the world is now proceeding towards the third wave of the pandemic^[13]^. In several countries, the third wave has already started and in many other countries it is yet to start. The aim of this study was to learn from the nature of initial two waves and to better prepare for the next unforeseen third wave.

In this article, we have analyzed the available data from those countries where the second or third waves of the pandemic has already been observed. Our aim was to use the result to predict the situations of those countries where the second or third wave is yet to strike so that preventive measures can be taken within the available time. We specially applied our obtained result to predict the situation of India. We tried to estimate the available time and probable number of persons to be infected so that the required precautions can be taken to avoid worst situation.

## 2. Methods

### Mathematical Analysis

We have used simple numerical interpolation method to derive our experimental formulae, containing some arbitrary constants or parameters. To obtain the values of those arbitrary parameters, basic concepts of calculus has been used and to determine their numerical values, statistical data has been used successfully. The details of mathematical interpretation, parameters and the constants used for this study have been described in the **Supplemental Method**. The figures were obtained using the MATLAB™.

### Ethical Approval and Data Source

The data for the mathematical analysis was obtained from free reference websites, such as Worldometer and/or Wikipedia. Thus, this analysis does not require any ethical approval.

## 3. Experiment

### 3.1 Nature of three waves of COVID-19 pandemic in different countries

We have chosen fifteen countries from different continents. Eight among them have already witnessed the third wave. In the other, the second wave has been observed till the time of experiment. The choice of the countries was completely arbitrary, based on population size (Brazil and Nepal), density (Bangladesh and Australia), weather (Russia and Colombia)^[14-15]^, and health infrastructure (Japan and Nigeria)^[16-17]^. The reason for taking such countries with contrasting natural and economic conditions was that we wanted to include the effect of weather, temperature, humidity, latitude^[5-12,19]^ on the rate of infection. The data was recorded until 25^th^ April 2021^[8]^. The starting date of first wave of a country was the day when first case reported. The ending date of first wave was considered on the day between first and second wave when the number of daily reported cases was minimum^[40]^. Similarly, the ending date of second wave was considered on the day between second and third wave when the number of daily reported cases was lowest^[40]^. The starting date of second wave was the ending date of first wave^[40]^ and in the same way the starting date of third wave was the ending date of second wave^[40]^. The date was given in the format DD/MM/YY (**Supplemental Table 1**).

### 3.2 Comparison of Rate of Infection and Rate of Death of three waves

1. From the **Supplemental Table 2**, it was observed that in the second wave, the rate of infection was **3.78 times** more than that of first wave. This indicated that the SARS-CoV2 mutated itself during the second wave^[19]^ in such a way that it could survive easily in the different conditions of a country^[19]^.
2. Though the average value of death rate showed that it was almost equal in first and second waves, but the virulent strength of the virus, in the second wave was less than that of first wave^[19,44-45]^. This was because in most of the countries, the rate of death in second wave was less than that of first wave. There were a few countries where the converse was true. However, the second wave was not still over in all countries and so the reality might change a little bit from our assumption.
3. The third wave was found to be more dangerous^[34,42]^. The rate of infection was **1.7 times** more than that of second wave and **6.23 times** more than the first wave. Although the death rate was **1.21 times** more than second wave, it was not as fatal as the first wave (at least **0.46 times** less than the first wave) (**Supplemental Table 2**).

It was also necessary to mention that the countries where the third wave started are not over yet. In most of the countries, the third wave has been found to be in the middle or in the early stage. So, we assumed that the rate of infection for third wave was much more than **6.23** times the first wave. Japan was such a country, where the third wave was almost over within the date of our experiment and the fourth wave was about to start. There the rate of infection was almost **20 times** more than that of first wave^[36]^.

### 3.3 Comparison of highest infections of three waves

Here we tried to estimate the highest number of infected people in a single day in each waves of infection and their comparative ratios.

1. From the **Supplemental Table 3** it was obvious that in the second wave, the highest number of persons, infected in a single day, was almost **2.85 times** more than that of first wave.
2. Similarly in the third wave, that was almost **1.778 times** more than second wave and **5.137 times** more than first wave.

### 3.4 Duration of three waves

1. In the **Supplemental Table 4**, it was found that the average duration of the first wave was around **193.4 days** *i.e*., **6 months** and **13 days**. Similarly, the average duration of the second wave was around **163.1 days** *i.e*., **5 months** and **13 days**.
2. It took about **187.3 days** *i.e*., **6 months** and **7 days** for the second wave to reach at its maximum value from the day when the first wave reached to its maximum value. Similarly, after the day when the maximum number of cases of second wave was obtained, almost **104.625 days** *i.e*., **3 months** and **15 days** were required for the third wave to reach to its maximum (**Supplemental Table 4**). So, the third wave has been striking almost just after the second wave.
3. It could be assumed that there was a time between two consecutive waves when the infection maintained a moderate rate. Between first and second wave, it was about **93.6 days** *i.e*., **3 months** and **4 days** and between second and third wave, it was almost **52.5 days** *i.e*., **1 month** and **23 days (Supplemental Table 4)**. This period was very crucial^[41]^. All the countries who were able to control the rate of infection, used this time frame to retard the infection rate^[41]^. Vaccination, regular sanitization, partial lockdown at that time could be helpful^[18,31,41,42]^.

### 3.5 Relation of population with duration of two waves

Now we considered those nine countries where the second wave was assumed to be completed till the time of experiment. We arranged them in decreasing order of population for convenience. If the population of the country was more than 50 million, we considered it high, otherwise low.

The population increases where the first wave usually remained longer than the second wave. Converse was true for the countries with low population. In those countries, the second wave was longer (**Supplemental Table 5**).

## 4. Results

### 4.1 Case Study on Ukraine

To support our hypothesis, we took the example of **Ukraine**^[23-24]^.

Total population: 43515909

Date of first case, reported: 03/03/2020.

Date of highest infection, reported in one day, in the first wave: 28/11/2020.

Date of highest infection, reported in one day, in the second wave: 03/04/2021.

Highest infection, reported in one day, in the first wave: 16294.

Highest infection, reported in one day, in the second wave: 20341.

Half of highest infection, reported in one day, in the first wave: 8312.

Half of highest infection, reported in one day, in the second wave: 10155.

Date of half of highest infection, reported in one day, in the first wave: 30/10/2020.

Date of half of highest infection, reported in one day, in the second wave: 05/03/2021.

Hence for **first wave**: k_1_ = 16294

> k_2_= 0.00077
>
> k_3_= 270

For **second wave**: k_1_ = 20341

> k_2_= 0.000884
>
> k_3_= 396

We took the number of infected people after an interval of 15 days (**Supplemental Table 6**).

Now the total number of infected people in Ukraine till 25/04/2021 was **2025271**^[23]^. And our formula (7) (**Supplemental Method**) gave-Number of infected people till 25/04/2021 was **2057485** which was close to the actual figure, with an error of just 1.5% (**Fig 1**).

**Figure 1:**
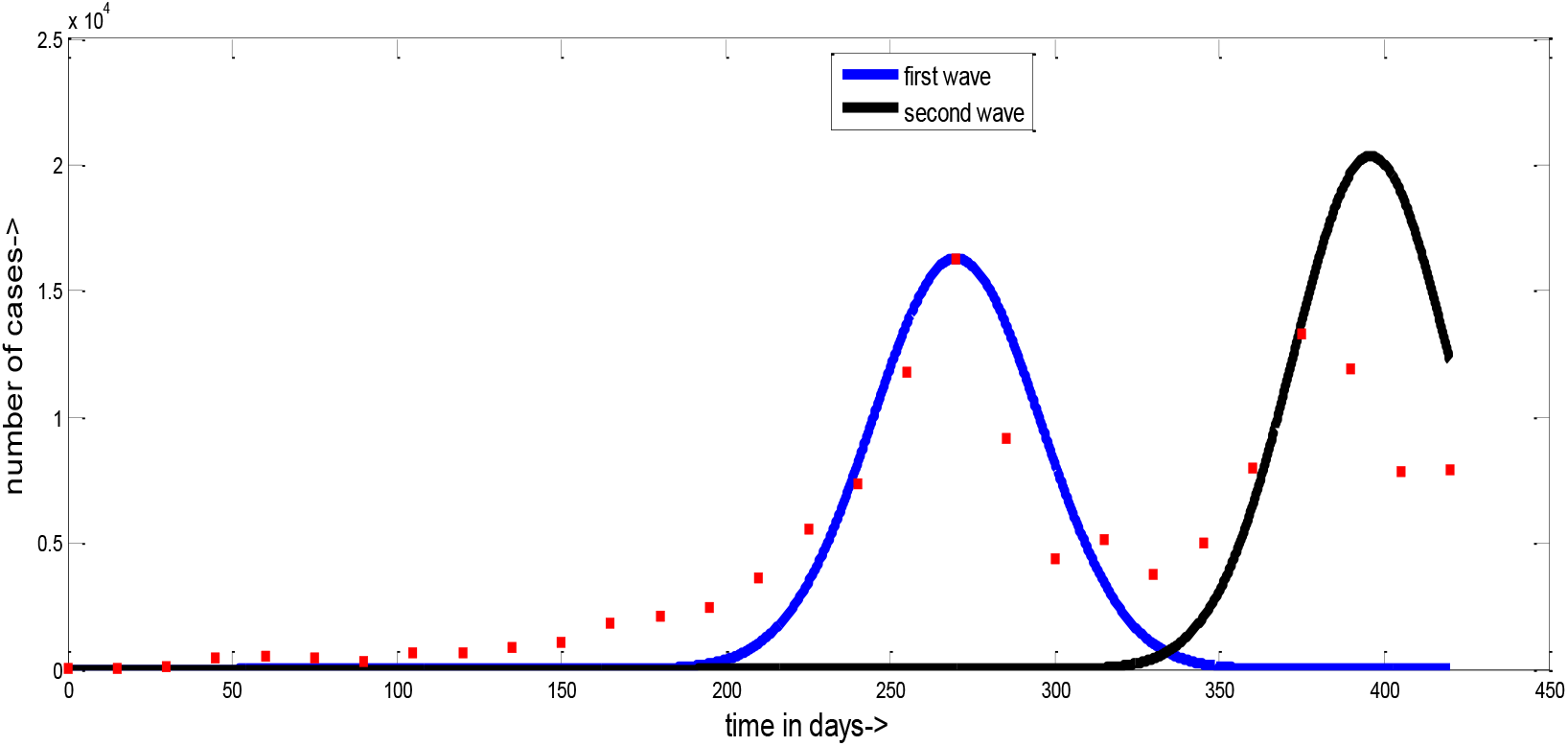
Predicted and Actual COVID-19 Waves of Ukraine. The plot showing the actual COVID-19 situation and our predicted situation of Ukraine. The number of days from starting of the pandemic is plotted along x axis and the number of cases along y axis. The red dots represent the actual cases and lines represent our prediction.

### 4.2 Case Study on Panama

Now we demonstrated how to apply our model to predict the situations of those countries where the second wave or third wave is yet to strike. For example, we took the **Panama** country^[25-26]^.

Total population: 4314767

Date of first case, reported: 09/03/2020.

Date of highest infection, reported in one day, in the first wave: 13/07/2020.

Highest infection, reported in one day, in the first wave: 1540.

Half of highest infection, reported in one day, in the first wave: 754.

Date of half of highest infection, reported in one day, in the first wave: 18/06/2020.

Hence for **first wave**: k_1_ = 1540

> k_2_= 0.00111
>
> k_3_= 126

If we considered the highest single day reported cases for second wave, was 2.85 times than that of first wave, then the highest single day case for second wave would be 1540*2.85 = 4389.

So, for **second wave**, k_1_ = 4389

The maximum number of reported cases should appear after about 6 months and 7 days from 13/07/2020. So, the maximum cases of second wave should appear on 16/01/2021. But since the population of panama is very low, we assumed the highest infection would occur around 31/12/2020.

Hence for **second wave**, k_3_ = 296

We apply formula (5) (**Supplemental Method**) to determine k_2_. The second wave was assumed to start when the number of cases is minimum between first and second wave^[40]^ *i.e*., we assumed the second wave to start from 04/11/2020 and on that day, the number of cases would be 432. So, on 04/11/2020, t =126+114 = 240. Hence from (5)

k_2_ = ln (4389/432)/(240-296)^2^ = 0.00074

We took the number of infected people after an interval of 15 days (**Supplemental Table 7**).

Now the total number of infected people in Panama till 25/04/2021 was **362967**^[25]^. And our formula (7) (**Supplemental Method**) gave-Number of infected people till 25/04/2021 was **363436** which was also close to the actual figure, with an error of just 0.12% (**Fig 2**).

**Figure 2:**
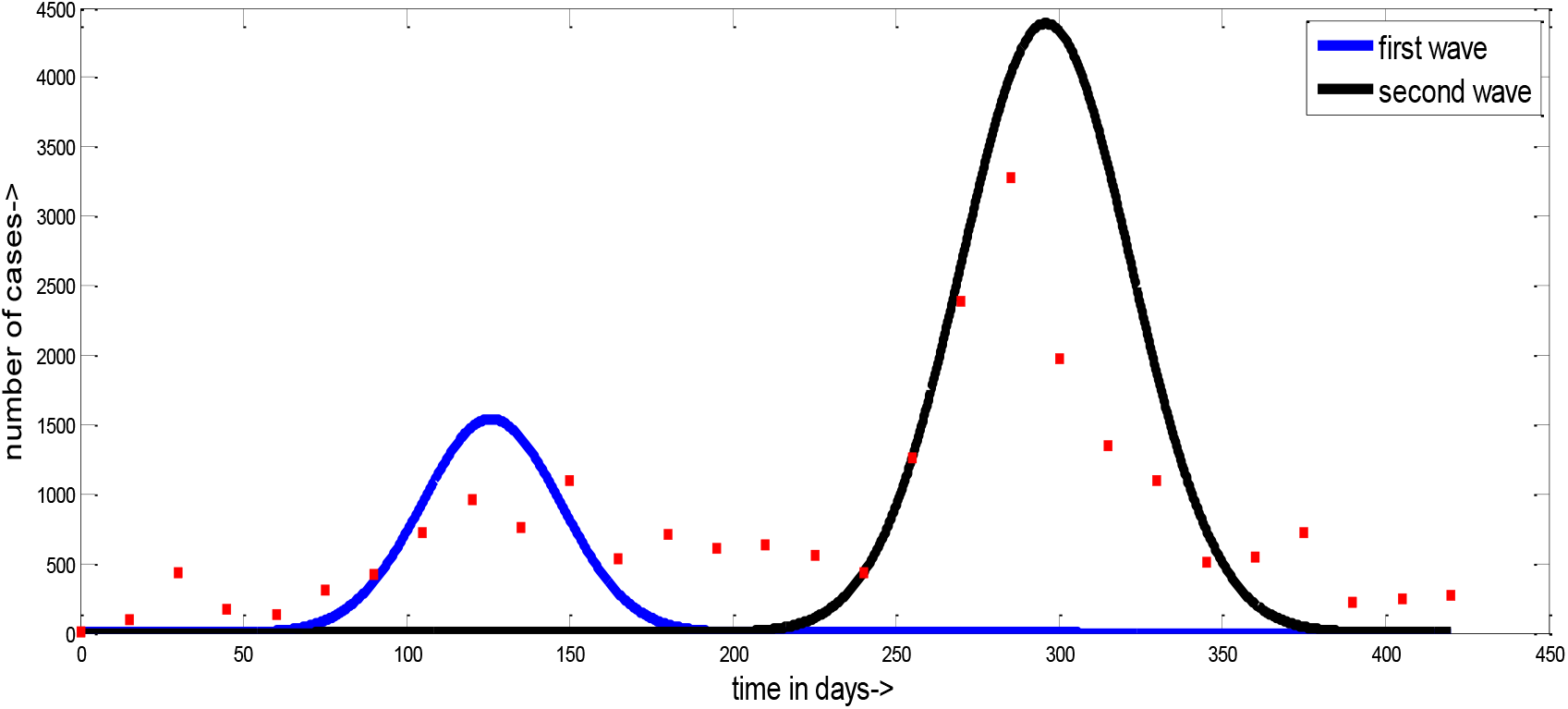
Predicted and Actual COVID-19 Waves of Panama. This is the figure of actual COVID-19 situation and our predicted situation of Panama. The number of Days from starting of the pandemic is plotted along x axis and the number of cases along y axis. The red dots represent the actual cases and lines represent our prediction.

### 4.3 Situation in India

Now coming to the situation in India; India is a country with huge population density^[29]^. The first wave of the pandemic peaked during September 2020 and the second wave has been surging from the beginning of April 2021^[27-28]^.

Total population: 1380004385

Date of first case, reported: 30/01/2020.

Date of highest infection, reported in one day, in the first wave: 16/09/2020.

Highest infection, reported in one day, in the first wave: 97859.

Half of highest infection, reported in one day, in the first wave: 49632.

Date of half of highest infection, reported in one day, in the first wave: 28/07/2020.

Hence for **first wave**: k_1_ = 97859

> k_2_= 0.0002772589
>
> k_3_= 230

If we considered the highest single day reported cases for second wave, was 2.85 times than that of first wave, then the highest single day case for second wave would be 97859*2.85 = 278898. But since the population of India is high, we consider the highest infected in one day in second wave was 350000.

So, for **second wave**, k_1_ = 350000

The maximum number of reported cases should appear after about 6 months and 7 days from 16/09/2020. So, the maximum case of second wave should appear on 28/03/2021. But since the population of India is very high^[29]^, wed assumed the highest infection would occur around 14/05/2021.

Hence for **second wave**, k_3_ = 469

We applied formula (5) (**Supplemental Method**) to determine k_2_. The second wave was assumed to be started when the number of cases was minimum between first and second wave^[40]^ *i.e*., we assumed the second wave to start from 01/02/2021 and on that day, the number of cases is 8587. So, on 01/02/2021, t = 368. Hence from (5),

k_2_= ln (350000/8587)/(368-469)^2^ = 0.0003634628

If we considered the highest single day reported cases for third wave, was 1.778 times than that of second wave, then the highest single day case for second wave would be 350000*1.778 = 622300.

So, for **third wave**, k_1_ = 622300

The maximum number of reported cases should appear after about 3 months and 15 days from 14/05/2021. So, the maximum case of third wave should appear on 28/08/2021. But since the population of India is very high^[29]^, we assumed the highest infection will occur around 05/09/2021.

Hence for **third wave**, k_3_ = 584

We applied formula (5) (**Supplemental Method**) to determine k_2_. The third wave was assumed to start when the number of cases is minimum between second and third wave^[40]^ *i.e*., we assume the third wave to start from 30/07/2021 and on that day, the number of cases should be near to the number on 15/03/2021 *i.e*., around 24437. So, on 30/07/2021, t = 549. Hence from (5)

k_2_ = ln (622300/24437)/(549-584)^2^ = 0.0026427

We took the number of infected people after an interval of 15 days (**Supplemental Table 8**).

Now to calculate the total number of affected people, we applied Formula (7) (**Supplemental Method**) which gave an approximate value of total number of infected people after three waves which was around **63647579** (**Fig 3**).

**Figure 3:**
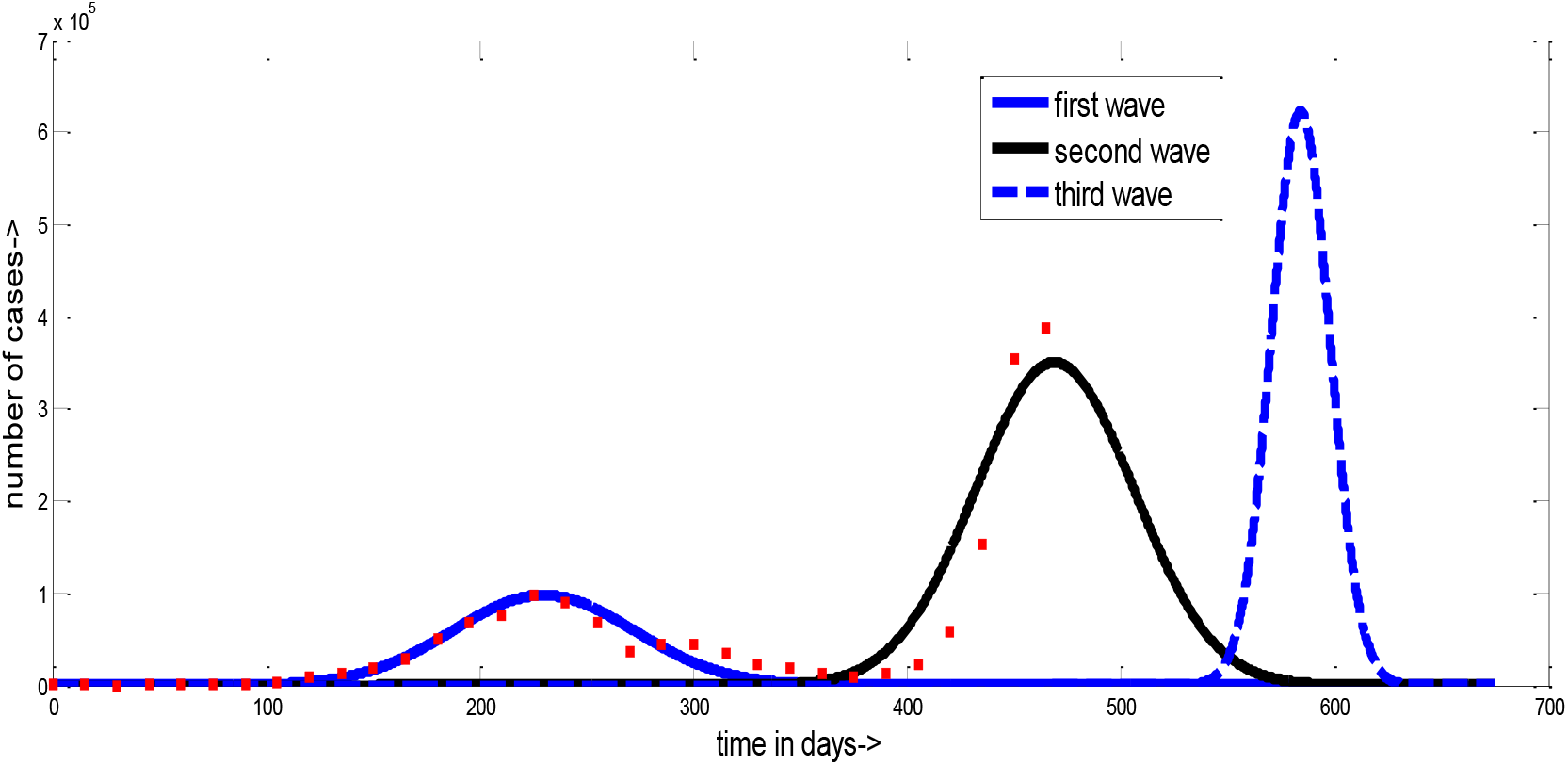
Predicted and Actual COVID-19 Waves of India. This is the figure of actual COVID-19 situation and our predicted situation of India. The number of days from starting of the pandemic is plotted along x axis and the number of cases along y axis. The red dots represent the actual cases and lines represent our prediction.

## 5 Discussion

Our investigation suggests that it is a high time for Indian Government to be prepared for the third wave. It is evident that the second and third wave was more dangerous than first wave of influenza pandemic (during 1818-1819)^[42]^, which was akin to the COVID-19 pandemic. We should not repeat the mistakes that we did in case of second wave^[41,44]^. Assuming the second wave would peak around mid-May, the third wave is predicted to peak around early September. Therefore, there is just 3-4 months of time in hand. According to reports, it is necessary to vaccinate **60-70%** of the total population within this predicted timeframe^[41,44,46]^. Vaccination and partial lockdown would reduce the infection almost half of the prediction^[43]^. We also predict that the curve will be much steep. However, the interval of the third wave is going to be much shorter: roughly meaning it being much easier for the situation to get out of control.

Countries like Australia, South Africa have become successful to control the infection after the second wave^[41]^, which shows that, though herculean, the task is not impossible. It would be helpful to put a lockdown of about one month in the month of July, when the second wave will be about to end^[31]^.

Besides this, we must develop the health infrastructure, increase the speed of vaccination^[41]^ and the number of tests. Masking, sanitization and social distancing should be made compulsory. The common people must show more responsibility and willingness to follow protocol.^[18]^. All kinds of meeting, social, political, or religious gathering should be strictly banned^[18]^ or conducted through virtual mode^[42]^.

The daily number of infected cases has already crossed our prediction. In the first wave (i.e., till 01/02/2021) total number of cases in India, was 10.7 million and our prediction shows almost 65 million people can be infected in this pandemic. These are only official figures with the true value likely to be higher due to less reporting or there being patients who have not been tested. However, no one would be happier than us, if our prediction goes wrong and we keep the infection under control. So, the common people as well as Government must be strict now, following the successful footsteps of pioneering countries^[41]^ (like New Zealand^[38,41]^, Australia^[39,41]^) in controlling the pandemic.

## 6 Conclusion

The population of each country was of 2020^[30]^ and it was considered constant to avoid difficulties in calculation. If the change of population was considered, then the result would differ a little from our findings. It is also important to mention that the process, we applied here, can be fruitful to estimate the probable number of active cases and deaths, from the early stage of a pandemic so that preventive measures and precautions can be taken according to the prediction.

## Data Availability

The data for the mathematical analysis was obtained from free reference websites, such as Worldometer and/or Wikipedia. Therefore, it is a secondary source data.

https://www.worldometers.info/coronavirus/

## 7 Acknowledgements

We thank all the frontline COVID-19 warriors for saving the mankind during this pandemic.

## 8 Author Contributions

T.S., A.B. and A.G. conceptualized the project, A.B. and S.R. designed the experiments, A.B. and S.R. performed the experiments and analyzed the data. A.G., A.B., S.R., and T.S. wrote the manuscript. T.S. and A.G. supervised the project.

## Reference

(1) World Health Organization, 2020. Origin of SARS-CoV-2, 26 March 2020 (No. WHO/2019-nCoV/FAQ/Virus_origin/2020.1). World Health Organization.

(2) Ye, Z.W., Yuan, S., Yuen, K.S., Fung, S.Y., Chan, C.P. and Jin, D.Y., 2020. Zoonotic origins of human coronaviruses. International journal of biological sciences, 16(10), p.1686.

(3) Poutanen, S.M., 2018. Human coronaviruses. Principles and Practice of Pediatric Infectious Diseases, p.1148.

(4) Zhu, Z., Lian, X., Su, X., Wu, W., Marraro, G.A. and Zeng, Y., 2020. From SARS and MERS to COVID-19: a brief summary and comparison of severe acute respiratory infections caused by three highly pathogenic human coronaviruses. Respiratory research, 21(1), pp.1–14.

(5) Brassey, J., Heneghan, C., Mahtani, K.R. and Aronson, J.K., 2020. Do weather conditions influence the transmission of the coronavirus (SARS-CoV-2). CEMB, www.cemb.net/oxford-COVID-19.

(6) Lin, K.U.N., Fong, D.Y.T., Zhu, B. and Karlberg, J., 2006. Environmental factors on the SARS epidemic: air temperature, passage of time and multiplicative effect of hospital infection. Epidemiology & Infection, 134(2), pp.223–230.

(7) Chan, K.H., Peiris, J.M., Lam, S.Y., Poon, L.L.M., Yuen, K.Y. and Seto, W.H., 2011. The effects of temperature and relative humidity on the viability of the SARS coronavirus. Advances in virology, 2011.

(8) Burra, P., Soto-Díaz, K., Chalen, I., Gonzalez-Ricon, R.J., Istanto, D. and Caetano-Anollés, G., 2021. Temperature and Latitude Correlate with SARS-CoV-2 Epidemiological Variables but not with Genomic Change Worldwide. Evolutionary Bioinformatics, 17, p.1176934321989695.

(9) Sajadi, M.M., Habibzadeh, P., Vintzileos, A., Shokouhi, S., Miralles-Wilhelm, F. and Amoroso, A., 2020. Temperature, humidity, and latitude analysis to estimate potential spread and seasonality of coronavirus disease 2019 (COVID-19). JAMA network open, 3(6), pp.e2011834–e2011834.

(10) Chennakesavulu, K. and Reddy, G.R., 2020. The effect of latitude and PM2. 5 on spreading of SARS-CoV-2 in tropical and temperate zone countries. Environmental Pollution, 266, p.115176.

(11) Whittemore, P.B., 2020. COVID-19 fatalities, latitude, sunlight, and vitamin D. American journal of infection control, 48(9), pp.1042–1044.

(12) Salman, A.D., AL-farttoosi, H.A.D. and Kadhim, A.J., 2020, November. Study impact the latitude on COVID-19 spread virus by data mining algorithm. In Journal of Physics: Conference Series (Vol. 1664, No. 1, p. 012109). IOP Publishing.

(13) URL-1: https://www.worldometers.info/coronavirus/

(14) URL-2: https://www.nationsencyclopedia.com/Europe/Russia-CLIMATE.html

(15) URL-3: https://www.nationsencyclopedia.com/Americas/Colombia-CLIMATE.html

(16) URL-4: https://www.nationsencyclopedia.com/Asia-and-Oceania/Japan-HEALTH.html

(17) URL-5: https://www.nationsencyclopedia.com/Africa/Nigeria-HEALTH.html

(18) Singley, A. and Callender Highlander, H., 2020. A Mathematical Model for the Effect of Social Distancing on the Spread of COVID-19. Spora: A Journal of Biomathematics, 6(1), pp.40–51.

(19) Anthony, S.J., Johnson, C.K., Greig, D.J., Kramer, S., Che, X., Wells, H., Hicks, A.L., Joly, D.O., Wolfe, N.D., Daszak, P. and Karesh, W., 2017. Global patterns in coronavirus diversity. Virus evolution, 3(1).

(20) URL-6: https://en.wikipedia.org/wiki/Probability_density_function

(21) URL-7: https://en.wikipedia.org/wiki/Normal_distribution

(22) URL-8: https://en.wikipedia.org/wiki/Simpson%27s_rule#:∼:text=Simpson’s%203%2F8%20rule%2C%20also,polynomial%20up%20to%20cubic%20degree.

(23) URL-9: https://www.worldometers.info/coronavirus/country/ukraine/

(24) URL-10: https://en.wikipedia.org/wiki/COVID-19_pandemic_in_Ukraine

(25) URL-11: https://www.worldometers.info/coronavirus/country/panama/

(26) URL-12: https://en.wikipedia.org/wiki/COVID-19_pandemic_in_Panama

(27) URL-13: https://www.worldometers.info/coronavirus/country/india/

(28) URL-14: https://en.wikipedia.org/wiki/COVID-19_pandemic_in_India

(29) URL-15: https://www.worldometers.info/world-population/india-population/

(30) URL-16: https://www.worldometers.info/population/

(31) Biswas, S.K., Ghosh, J.K., Sarkar, S. and Ghosh, U., 2020. COVID-19 pandemic in India: a mathematical model study. Nonlinear dynamics, 102(1), pp.537–553.

(32) Ghosh, S., Samanta, G.P. and Mubayi, A., 2021. Comparison of Regression Approaches for Analyzing Survival Data in the Presence of Competing Risks. Letters in Biomathematics, 8(1), pp.29–47.

(33) Ghanam, R., Boone, E.L. and Abdel-Salam, A.S.G., 2020. Seird model for qatar COVID-19 outbreak: A case study. arXiv preprint 2005.12777.

(34) Seong, H., Hyun, H.J., Yun, J.G., Noh, J.Y., Cheong, H.J., Kim, W.J. and Song, J.Y., 2021. Comparison of the second and third waves of the COVID-19 pandemic in South Korea: Importance of early public health intervention. International Journal of Infectious Diseases, 104, pp.742–745.

(35) Post, L.A., Benishay, E.T., Moss, C.B., Murphy, R.L., Achenbach, C.J., Ison, M.G., Resnick, D., Singh, L.N., White, J., Chaudhury, A.S. and Boctor, M.J., 2021. Surveillance metrics of SARS-CoV-2 transmission in central Asia: longitudinal trend analysis. Journal of medical Internet research, 23(2), p.e25799.

(36) URL-17: https://www.worldometers.info/coronavirus/country/japan/

(37) URL-18: https://en.wikipedia.org/wiki/Proof_by_contradiction

(38) URL-19: https://www.worldometers.info/coronavirus/country/new-zealand/

(39) URL-20: https://www.worldometers.info/coronavirus/country/australia/

(40) Xiang, N., Iuliano, A.D., Zhang, Y., Ren, R., Geng, X., Ye, B., Tu, W., ao Li, C., Lv, Y., Yang, M. and Zhao, J., 2016. Comparison of the first three waves of avian influenza A (H7N9) virus circulation in the mainland of the People’s Republic of China. BMC infectious diseases, 16(1), pp.1–12.

(41) Graichen, H., 2021. What is the difference between the first and the second/third wave of COVID-19?–German perspective.

(42) Paulo, A.C., Correia-Neves, M., Domingos, T., Murta, A.G. and Pedrosa, J., 2010. Influenza infectious dose may explain the high mortality of the second and third wave of 1918–1919 influenza pandemic. PLoS One, 5(7), p.e11655.

(43) URL-21: https://www.nature.com/articles/d41586-020-00502-w

(44) URL-22: https://www.orfonline.org/expert-speak/second-wave-COVID-19-causes-solutions/

(45) URL-23: https://science.thewire.in/health/sars-cov-2-variants-b117-b1617-india-second-wave-uncertain-future/

(46) URL-24: https://www.who.int/news-room/q-a-detail/herd-immunity-lockdowns-and-COVID-19?gclid=CjwKCAjwm7mEBhBsEiwA_of-TCu-qBllk7fkColTirzjCycCNNc5arV_PDWst6hscs-l0JTRi47HsRoC2bUQAvD_BwE#

