## Supplemental Information for "Dynamics of the Third wave, modelling COVID-19 pandemic with an outlook towards India"

*Corresponding Author

*Supplemental Table 1. Nature of three waves of COVID-19 pandemic in different countries[13]*

| Country | Population | First wave | | | | Second wave | | | | Third wave | | | |
|---|---|---|---|---|---|---|---|---|---|---|---|---|---|
| | | Starts on | Highest infection | Total infected | ROI* | Starts on | Highest infection | Total infected | ROI* | Starts on | Highest infection | Total infected | ROI* |
| | Density# | Closes on | Date | Death | ROD** | Closes on | Date | Death | ROD** | Close on | Date | Death | ROD** |
| Colombia | 51309459 | 6/03/20 | 13056 | 818203 | 1.594644 | 28/09/20 | 21078 | 1460653 | 2.846752 | 8/03/21 | 19925 | 478413 | 0.932407 |
| | 46 | 28/09/20 | 19/08/20 | 25641 | 3.133819 | 8/03/21 | 15/01/21 | 34957 | 2.393245 | | 23/04/21 | 10288 | 2.150443 |
| Canada | 38002618 | 27/01/20 | 2760 | 105536 | 0.277707 | 5/07/20 | 11383 | 721388 | 1.898259 | 15/02/21 | 11041 | 345080 | 0.908043 |
| | 4 | 5/07/20 | 3/05/20 | 8684 | 8.228472 | 15/02/21 | 3/01/21 | 12627 | 1.750376 | | 3/04/21 | 2616 | 0.758085 |
| Mexico | 129998266 | 28/02/20 | 9866 | 817503 | 0.628857 | 12/10/20 | 22339 | 1505927 | 1.158421 | | | | |
| | 66 | 12/10/20 | 2/08/20 | 93899 | 11.48607 | | 22/01/21 | 120605 | 8.008688 | | | | |
| Brazil | 213749436 | 25/02/20 | 70869 | 5749007 | 2.689601 | 2/11/20 | 87134 | 4085506 | 1.911353 | 14/02/21 | 97586 | 4473702 | 2.092966 |
| | 25 | 2/11/20 | 29/07/20 | 160272 | 2.787821 | 14/02/21 | 7/01/21 | 79022 | 1.934204 | | 25/03/21 | 150315 | 3.359969 |
| England | 67886011 | 22/01/20 | 7846 | 286077 | 0.421408 | 6/07/20 | 33409 | 1249901 | 1.841176 | 24/11/20 | 67928 | 2867192 | 4.223539 |
| | 281 | 6/07/20 | 10/04/20 | 40696 | 14.22554 | 24/11/20 | 12/11/20 | 15211 | 1.216976 | | 8/01/21 | 71510 | 2.494078 |
| Spain | 46769469 | 31/01/20 | 10401 | 250177 | 0.534915 | 11/06/20 | 22766 | 1466098 | 3.134733 | 6/12/20 | 34232 | 1752342 | 3.746765 |
| | 94 | 11/06/20 | 20/03/20 | 29288 | 11.70691 | 6/12/20 | 27/10/20 | 17226 | 1.174956 | | 15/01/21 | 31077 | 1.773455 |
| Nigeria | 210134738 | 27/02/20 | 790 | 59345 | 0.028241 | 4/10/20 | 2314 | 105339 | 0.050129 | | | | |
| | 226 | 4/10/20 | 1/07/20 | 1113 | 1.875474 | | 22/01/21 | 948 | 0.899952 | | | | |
| South Africa | 59896382 | 5/03/20 | 13944 | 693359 | 1.157598 | 12/10/20 | 21980 | 877989 | 1.465846 | | | | |
| | 49 | 12/10/20 | 24/07/20 | 18236 | 2.630095 | | 8/01/20 | 35889 | 4.087636 | | | | |
| Turkey | 85052838 | 11/03/20 | 13976 | 604943 | 0.711255 | 22/07/20 | 33198 | 1819385 | 2.139123 | 23/01/21 | 63082 | 2167088 | 2.547931 |
| | 110 | 22/07/20 | 11/04/20 | 5545 | 0.916615 | 23/01/21 | 8/12/20 | 15548 | 0.854575 | | 16/04/21 | 16918 | 0.780679 |
| Russia | 145983862 | 31/01/20 | 11656 | 970865 | 0.66505 | 26/08/20 | 29935 | 3782924 | 2.591331 | | | | |
| | 9 | 26/08/20 | 11/05/20 | 16683 | 1.718365 | | 24/12/20 | 91217 | 2.411283 | | | | |
| Nepal | 29547776 | 23/01/20 | 740 | 17061 | 0.05774 | 14/07/20 | 5743 | 257320 | 0.870861 | 7/03/21 | | | |
| | 203 | 14/07/20 | 3/07/20 | 45 | 0.263759 | 7/03/21 | 21/10/20 | 2965 | 1.152262 | | | | |
| Bangladesh | 165980237 | 8/03/20 | 4019 | 540266 | 0.3255 | 13/02/21 | 7626 | 688374 | 0.414732 | | | | |
| | 1265 | 13/02/21 | 2/07/20 | 8266 | 1.529987 | | 7/04/21 | 2686 | 0.390195 | | | | |
| Japan | 126170451 | 16/01/20 | 743 | 16804 | 0.013318 | 30/05/20 | 1998 | 63237 | 0.05012 | 24/09/20 | 7855 | 360630 | 0.285828 |
| | 347 | 30/05/20 | 11/04/20 | 886 | 5.272554 | 24/09/20 | 3/08/21 | 634 | 1.002578 | 9/03/21 | 9/01/21 | 6779 | 1.879766 |
| Iran | 84837813 | 19/02/20 | 3574 | 375212 | 0.44227 | 31/08/20 | 14051 | 825253 | 0.972742 | 27/12/20 | 25582 | 1176574 | 1.386851 |
| | 52 | 31/08/20 | 4/06/20 | 21571 | 5.749017 | 27/12/20 | 27/11/20 | 33122 | 4.013557 | | 14/04/21 | 14427 | 1.226187 |
| Australia | 25732506 | 25/01/20 | 537 | 7255 | 0.028194 | 6/06/20 | 721 | 22333 | 0.086789 | | | | |
| | 3 | 6/06/20 | 22/03/20 | 102 | 1.405927 | | 30/07/20 | 808 | 3.617965 | | | | |

***ROI =Rate of Infection given in percentage [31]; **ROD = Rate of Death given in percentage [31]; #Density is given per square kilometer**

### *Supplemental Table 2. Comparison of Rate of Infection and Rate of Death of three waves*

| **Country** | **First wave** | **Second wave** | **Third wave** | **$I_2/I_1$** | **$I_3/I_2$** | **$I_3/I_1$** |
|---|---|---|---|---|---|---|
| | **Rate of Infection($I_1$)** | **Rate of Infection($I_2$)** | **Rate of Infection($I_3$)** | | | |
| | **Rate of Death($D_1$)** | **Rate of Death($D_3$)** | **Rate of Death($D_3$)** | **$D_2/D_1$** | **$D_3/D_2$** | **$D_3/D_1$** |
| Colombia | 1.594644 | 2.846752 | 0.932401 | 1.785196 | 0.327534 | 0.584712 |
| | 3.133819 | 2.393245 | 2.150443 | 0.763683 | 0.898547 | 0.686205 |
| Canada | 0.277707 | 1.898259 | 0.908043 | 6.835474 | 0.478356 | 3.269788 |
| | 8.228472 | 1.750376 | 0.758085 | 0.212722 | 0.433098 | 0.09213 |
| Mexico | 0.628857 | 1.158421 | | 1.842106 | | |
| | 11.48607 | 8.008688 | | 0.697252 | | |
| Brazil | 2.689601 | 1.911353 | 2.092966 | 0.710646 | 1.095018 | 0.77817 |
| | 2.787821 | 1.934204 | 3.359969 | 0.693805 | 1.737133 | 1.205231 |
| England | 0.421408 | 1.841176 | 4.223539 | 4.369105 | 2.293935 | 10.022446 |
| | 14.22554 | 1.216976 | 2.494078 | 0.085549 | 2.049406 | 0.175324 |
| Spain | 0.534915 | 3.134733 | 3.746765 | 5.860245 | 1.195242 | 7.004412 |
| | 11.70691 | 1.174956 | 1.773455 | 0.100364 | 1.50938 | 0.151488 |
| Nigeria | 0.028241 | 0.050129 | | 1.775043 | | |
| | 1.875474 | 0.899952 | | 0.479853 | | |
| South Africa | 1.157598 | 1.465846 | | 1.266282 | | |
| | 2.630095 | 4.087636 | | 1.554178 | | |
| Turkey | 0.711255 | 2.139123 | 2.547931 | 3.007533 | 1.9111 | 3.582303 |
| | 0.916615 | 0.854575 | 0.780679 | 0.932316 | 0.913529 | 0.851698 |
| Russia | 0.66505 | 2.591331 | | 3.896445 | | |
| | 1.718365 | 2.411283 | | 1.403243 | | |
| Nepal | 0.05774 | 0.870861 | | 15.082456 | | |
| | 0.263759 | 1.152262 | | 4.368617 | | |
| Bangladesh | 0.3255 | 0.414732 | | 1.274138 | | |
| | 1.529987 | 0.390195 | | 0.256179 | | |
| Japan | 0.013318 | 0.05012 | 0.285828 | 3.763328 | 5.702873 | 21.461781 |
| | 5.272554 | 1.002578 | 1.879766 | 0.19015 | 1.874932 | 0.356519 |
| Iran | 0.44227 | 0.972742 | 1.386851 | 2.19943 | 1.425718 | 3.135756 |
| | 5.749017 | 4.013557 | 1.226187 | 0.698129 | 0.305511 | 0.213286 |
| Australia | 0.028194 | 0.086789 | | 3.078279 | | |
| | 1.405927 | 3.617965 | | 2.573366 | | |
| ***Average*** | | | | **3.7830472** | **1.713722** | **6.229921** |
| | | | | **1.000627** | **1.215192** | **0.466485** |

### *Supplemental Table 3. Comparison of highest infections of three waves*

| **Country** | **Highest infection in First wave ($H_1$)** | **Highest infection in Second wave ($H_2$)** | **Highest infection in Third wave ($H_3$)** | **$H_2/H_1$** | **$H_3/H_2$** | **$H_3/H_1$** |
|---|---|---|---|---|---|---|
| **Colombia** | 13056 | 21078 | 19925 | 1.61443 | 0.945298 | 1.526118 |
| **Canada** | 2760 | 11383 | 11041 | 4.124275 | 0.969955 | 4.000362 |
| **Mexico** | 9866 | 22339 | | 2.264241 | | |
| **Brazil** | 70869 | 87134 | 97586 | 1.229508 | 1.119953 | 1.376991 |
| **England** | 7846 | 33409 | 67928 | 4.258093 | 2.033225 | 8.65766 |
| **Spain** | 10401 | 22766 | 34232 | 2.188828 | 1.503646 | 3.291222 |
| **Nigeria** | 790 | 2314 | | 2.929114 | | |
| **South Africa** | 13944 | 21980 | | 1.576305 | | |
| **Turkey** | 13976 | 33198 | 63082 | 2.375358 | 1.900175 | 4.513595 |
| **Russia** | 11656 | 29935 | | 2.568205 | | |
| **Nepal** | 740 | 5743 | | 7.760811 | | |
| **Bangladesh** | 4019 | 7626 | | 1.897487 | | |
| **Japan** | 743 | 1998 | 7855 | 2.689098 | 3.93143 | 10.562005 |
| **Iran** | 3574 | 14051 | 25582 | 3.931449 | 1.820653 | 7.157806 |
| **Australia** | 537 | 721 | | 1.342644 | | |
| ***Average*** | | | | 2.85 | 1.778 | 5.137 |

### *Supplemental Table 4. Duration of three waves*

| **Country** | **Duration of First wave in days($D_1$)** | **Duration of Second wave in days($D_2$)** | **$D_1$+$D_2$** | **Time between highest cases of first and second wave($T_1$)** | **Time between highest cases of second and third wave($T_2$)** | **$T_3=T_1/2$ (in days)** | **$T_4=T_3/2$ (in days)** |
|---|---|---|---|---|---|---|---|
| **Colombia** | 206 | 159 | 365 | 149 | 97 | 75 | 49 |
| **Canada** | 160 | 225 | 385 | 245 | 91 | 123 | 46 |
| **Mexico** | 227 | | | 174 | | 87 | |
| **Brazil** | 251 | 104 | 355 | 162 | 78 | 81 | 39 |
| **England** | 166 | 141 | 307 | 216 | 57 | 108 | 29 |
| **Spain** | 132 | 178 | 310 | 217 | 88 | 108 | 44 |
| **Nigeria** | 220 | | | 206 | | 103 | |
| **South Africa** | 221 | | | 168 | | 84 | |
| **Turkey** | 133 | 184 | 317 | 241 | 130 | 120 | 65 |
| **Russia** | 208 | | | 226 | | 113 | |
| **Nepal** | 173 | 240 | 413 | 108 | | 54 | |
| **Bangladesh** | 340 | | | 279 | | 139 | |
| **Japan** | 136 | 118 | 253 | 115 | 159 | 58 | 80 |
| **Iran** | 195 | 119 | 314 | 173 | 137 | 86 | 68 |
| **Australia** | 133 | | | 130 | | 65 | |
| ***Average*** | 193.4 = 6 months 13 days | 163.1= 5 months 13 days | 335.4 = 11 months 5 days | 187.3 = 6 months 7 days | 104.625= 3 months 15 days | 93.6 = 3 months 4 days | 52.5 = 1 month 23 days |

### *Supplemental Table 5. Relation of population with duration of two waves*

| Country | Population | Duration of First wave in days($D_1$) | Duration of Second wave in days($D_2$) |
|---|---|---|---|
| **Brazil** | 213749436 | 251 | 104 |
| **Japan** | 126170451 | 136 | 118 |
| **Turkey** | 85052838 | 133 | 184 |
| **Iran** | 84837813 | 195 | 119 |
| **England** | 67886011 | 166 | 141 |
| **Colombia** | 51309459 | 206 | 159 |
| ***Average (from Table-4)*** | | 193.4 | 163.1 |
| **Spain** | 46769469 | 132 | 178 |
| **Canada** | 38002618 | 159 | 225 |
| **Nepal** | 29547776 | 173 | 240 |
| ***Average (from Table-4)*** | | 193.4 | 163.1 |

### *Supplemental Table 6. Case Study on Ukraine* [23-24]

| Date | Cases | Date | Cases | Date | Cases |
|---|---|---|---|---|---|
| 3/03/20 | 1 | 31/07/20 | 1090 | 28/12/20 | 4385 |
| 18/03/20 | 2 | 15/08/20 | 1847 | 12/01/21 | 5116 |
| 2/04/20 | 138 | 30/08/20 | 2096 | 27/01/21 | 3776 |
| 17/04/20 | 444 | 14/09/20 | 2462 | 11/02/21 | 5039 |
| 2/05/20 | 550 | 29/09/20 | 3627 | 26/02/21 | 8003 |
| 17/05/20 | 433 | 14/10/20 | 5590 | 13/03/21 | 13276 |
| 1/06/20 | 340 | 29/10/20 | 7342 | 28/03/21 | 7856 |
| 16/06/20 | 666 | 13/11/20 | 11787 | 12/04/21 | 7915 |
| 1/07/20 | 664 | 28/11/20 | 16294 | 25/04/21 | 7930 |
| 16/07/20 | 848 | 13/12/20 | 9176 | | |

### *Supplemental Table 7. Case Study on Panama* [25-26]

| Date | Cases | Date | Cases | Date | Cases |
|---|---|---|---|---|---|
| 9/03/20 | 1 | 6/08/20 | 1091 | 3/01/21 | 1972 |
| 24/03/20 | 98 | 21/08/20 | 537 | 18/01/21 | 1342 |
| 8/04/20 | 428 | 5/09/20 | 709 | 02/02/21 | 1098 |
| 23/04/20 | 171 | 20/09/20 | 602 | 17/02/21 | 504 |
| 8/05/20 | 137 | 5/10/20 | 633 | 4/03/21 | 540 |
| 23/05/20 | 310 | 20/10/20 | 558 | 19/03/21 | 715 |
| 7/06/20 | 421 | 4/11/20 | 432 | 3/04/21 | 223 |
| 22/06/20 | 722 | 19/11/20 | 1256 | 18/04/21 | 244 |
| 7/07/20 | 957 | 4/12/20 | 2388 | 25/04/21 | 271 |
| 22/07/20 | 753 | 19/12/20 | 3274 | | |

### *Supplemental Table 8. Situation in India*[27-28]

| Date | Cases | Date | Cases | Date | Cases |
|---|---|---|---|---|---|
| 30/01/2020 | 1 | 13/07/2020 | 28179 | 25/12/2020 | 22350 |
| 14/02/2020 | 2 | 28/07/2020 | 49632 | 09/01/2021 | 18820 |
| 29/02/2020 | 0 | 12/08/2020 | 67066 | 24/01/2021 | 13239 |
| 15/03/2020 | 26 | 27/08/2020 | 76826 | 08/02/2021 | 8947 |
| 30/03/2020 | 227 | 11/09/2020 | 97654 | 23/02/2021 | 13463 |
| 14/04/2020 | 1463 | 26/09/2020 | 89010 | 10/03/2021 | 22841 |
| 29/04/2020 | 1813 | 11/10/2020 | 67757 | 25/03/2021 | 59069 |
| 14/05/2020 | 3942 | 26/10/2020 | 36838 | 10/04/2021 | 152682 |
| 29/05/2020 | 8105 | 10/11/2020 | 44679 | 25/04/2021 | 354531 |
| 13/06/2020 | 12023 | 25/11/2020 | 44699 | | |
| 28/06/2020 | 19620 | 10/12/2020 | 34666 | | |

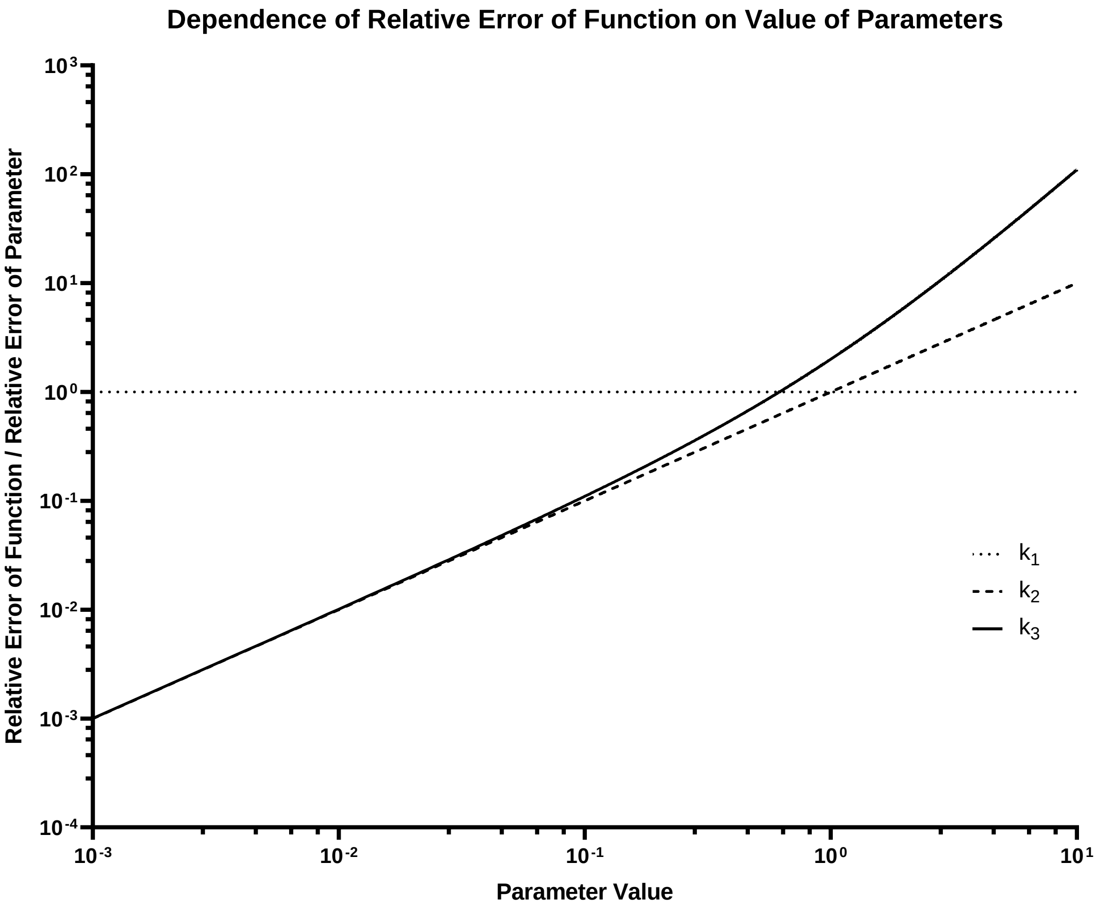


**Supplemental Figure 1:** Graph showing the variation of the relative error of the model function with respect to the relative errors in the parameters themselves based on variation in the value of the parameters.

***Supplemental Method:***

### 1. Mathematical Interpretation:

Considering any single wave (say, first wave) of any country, if we plotted ‘number of days from the starting of pandemic’ along x-axis and ‘number of infected persons on each day’ along y-axis, we found a ‘Bell-shaped’ curve like the graph of a probability density function [20] of Normal Distribution [21]. So, we tried to estimate it approximately by the following function-

$$f(t) = k_1 * \exp(- k_2 *(t - k_3)^2) \ldots\ldots\ldots\ldots\ldots\ldots(1)$$

where $k_1$ , $k_2$ , $k_3$ were nonzero (for the proof follow 4.2 Lemma ) arbitrary constants, ‘t’ was the time (measured in days) and $f(t)$ was the number of infected people at any time ‘t’ [31-33].

The arbitrary constants might depend on several factors like population [5-12, 31-33], density [5-12, 31-33], health infrastructure [5-12, 31-33], weather [5-12, 31-33] etc. So, for each country and each wave, $k_1$, $k_2$, $k_3$ were different and could be determined from the available data. We tried to explain an analytic method to find the value of $k_1$, $k_2$, $k_3$ so that our result best approximated the available data. Then we would try to predict the situations of India.

#### *1.1 Determining $k_1$, $k_2$, $k_3$:*

$$f(t) = k_1 * \exp(- k_2 *(t - k_3)^2 )$$

⇨ $df / dt = k_1 * \exp(- k_2 *(t - k_3)^2 )*(-2*k_2*(t-k_3))$ [differentiating both side w.r.t. ‘t’]

$$= 2*k_1*k_2* \exp(- k_2 *(t - k_3)^2 )*(k_3 - t) \ldots\ldots\ldots\ldots\ldots\ldots\ldots..(2)$$

Now at the extreme value of the function, $df / dt = 0$.

Now $df / dt = 0$

- ⇨ $2*k_1*k_2* \exp(- k_2 *(t - k_3)^2 )*(k_3 - t) = 0$
- ⇨ $k_3 - t = 0$ [since, $k_1$ , $k_2$ , $\exp(- k_2 *(t - k_3)^2 )$ could not be zero (follow 4.2 Lemma) ]
- ⇨ $t = k_3 \ldots\ldots\ldots\ldots\ldots\ldots\ldots.(3)$

Hence, we could conclude that the value of ‘t’ (time) when the curve reached to its maximum, giving the value of $k_3$.

Now let us put $t = k_3$ in (1) -

$$f(k_3) = k_1 * \exp(-k_2 * (k_3 - k_3)^2) = k_1 * 1 = k_1 \text{ ………………… } (4)$$

Hence the maximum value of the function was nothing but $k_1$. Thus, $k_1$ was also obtained.

Now taking log to both side of (1)

$$\ln(f(t)) = \ln(k_1 * \exp(-k_2 * (t - k_3)^2)) = \ln(k_1) - k_2 * (t - k_3)^2$$

- ⇨ $k_2 = (\ln(k_1) - \ln(f(t))) / (t-k_3)^2 = \ln(k_1/f(t)) / (t-k_3)^2$ …..………..…(5)

This was the general formula to determine $k_2$, taking the value of $f(t)$ at any time ‘t’ from the graph.

Suppose on $t = T$, $f(T) = (1/2)*(f(k_3)) = (1/2)*(k_1)$ [from (4)]

Then $(1/2)*k_1 = k_1 * \exp(-k_2 * (T - k_3)^2)$

- ⇨ $(1/2) = \exp(-k_2 * (T - k_3)^2)$
- ⇨ $\ln(1/2) = \ln(\exp(-k_2 * (T - k_3)^2))$

$$= -k_2 * (T - k_3)^2$$

- ⇨ $k_2 = (-\ln(1/2))/(T - k_3)^2 = \ln(2)/(T - k_3)^2$ ……………………(6)

So, if we took the value T from the graph, then the formula (6) could be used to determine $k_2$. However, to determine $k_2$ the value of $f(t)$ could be chosen at any value of ‘t’ and then using formula (5).

***1.2 Lemma:*** The constants $k_1$, $k_2$, $k_3$ were nonzero.

***Proof:*** We used the method of contradiction [37] to prove the Lemma.

If possible, let $k_1 = 0$.

Then $f(t) = 0 * \exp(-k_2 * (t - k_3)^2) = 0$, for all ‘t’

⇨ The number of infected people was zero forever- which was not true. Hence $k_1$ was nonzero.

Now suppose $k_2 = 0$. Then-

$f(t) = k_1 * \exp(-0 * (t - k_3)^2)$

$= k_1 * \exp(0)$

$= k_1 * 1 = k_1$

implying that the number of COVID-19 cases was same on each day which was also contradictory to the real scenario. So $k_2$ was also nonzero.

Finally, we had to prove $k_3$ was also nonzero. Now this was an obvious fact since $k_3$ was the number of days from the initiation of pandemic, when the infection reached to its maximum value.

#### *1.3 Error analysis of f(t):*

To understand the robustness of our model, we checked the error of the output function f(t) based on error of the input parameters, viz. $k_1$, $k_2$ and $k_3$. To do that, we partially differentiated f(t) with respect to $k_1$, $k_2$ and $k_3$ respectively.

$$\frac{\delta f}{\delta k_1} = e^{-k_2 \cdot (t-k_3)^2} = \frac{f}{k_1} \Rightarrow \frac{\delta f}{f} = \frac{\delta k_1}{k_1}$$

$$\frac{\delta f}{\delta k_2} = -k_1 \cdot (t-k_3)^2 \cdot e^{-k_2 \cdot (t-k_3)^2} \Rightarrow \frac{\delta f}{f} = -(t-k_3)^2 \cdot \delta k_2 = \mathrm{c} \cdot k_2 \cdot \frac{\delta k_2}{k_2} \propto k_2 \cdot \frac{\delta k_2}{k_2}$$

$$\frac{\delta f}{\delta k_3} = 2 \cdot k_1 \cdot k_2 \cdot (t-k_3) \cdot e^{-k_2 \cdot (t-k_3)^2} \Rightarrow \frac{\delta f}{f} = \mathrm{c} \cdot (\mathrm{t} - k_3) \cdot k_3 \cdot \frac{\delta k_3}{k_3} \propto (k_3 + {k_3}^2) \cdot \frac{\delta k_3}{k_3}$$

Thus, we could see that the function was not equally sensitive to errors in the three parameters. The function was less sensitive to errors in $k_1$ than $k_2$ and $k_3$, with it being more sensitive to $k_3$ than $k_2$ and so we need to be more careful in estimating these parameters (**Supplemental Fig 1**).

#### *1.4 Determining the total number of affected people:*

Suppose for a particular country, the value of $k_1$, $k_2$, $k_3$ were known for a particular wave. Then we could easily determine the total number of affected people in that wave by simple integration. If the wave was assumed to be ended in $t_1$ days, then P, the total number of affected people in $t_1$ days was-

$$P = \int_0^{t1} f\,(t)\,dt = k_1 \int_0^{t1} e^{-k2(t-k3)^{\wedge}2}\,dt \quad \ldots\ldots\ldots\ldots\ldots\ldots\ldots. (7)$$

Now this integral could not be expressed in terms of elementary functions [21]. So, any numerical method could be applied to perform the integration. It was convenient to use a c program as given below. However, one can use any software like MATLAB™, Mathematica etc. or any numerical method to perform the integration. Here we applied Simpson's Three Eighth rule [22] with the help of a simple c program.

#### *1.5 Simpson's three eighth rule (C program)*

```
#include<stdio.h>
#include<math.h>
float f(float k1,float k2,float k3,float t);
main()
{
	int i,n,j;
	float d,sum,r,k1,k2,k3,h,t0,t1,x[1000],T,y[10];
	printf("enter the value of k1:");
	scanf("%f",&k1);
	printf("enter the value of T to determine k2:");
	scanf("%f",&T);
	printf("enter the value of k3:");
	scanf("%f",&k3);
	k2=log(2)/pow((T-k3),2);
	printf("enter the lower limit of integration:");
	scanf("%f",&t0);
```

```
	printf("enter the upper limit of integration:");
	scanf("%f",&t1);
	printf("enter the number of subintervals");
	scanf("%d",&n);
	h=(t1-t0)/n;
	for(i=1;i<=(n+1);i++)
	{
		x[i]=t0+(i-1)*h;
	}
	for(i=1;i<=n;i++)
	{
		d=(x[i+1]-x[i])/3;
		for(j=1;j<=4;j++)
		{
			y[j]=x[i]+(j-1)*d;
		}

	sum=3*d*(f(k1,k2,k3,y[1])+3*f(k1,k2,k3,y[2])+3*f(k1,k2,k3,y[3])+f(k1,k2,k3,y[4]))/8;
		r=r+sum;
	}
	printf("\nthe value of the integration is : %f",r);
}
float f(float k1,float k2,float k3,float t)
{
	float r;
	r=k1*exp((-k2)*(pow((t-k3),2)));
	return(r);
}
```
